# Evaluating SARS-CoV-2 spike and nucleocapsid proteins as targets for IgG antibody detection in severe and mild COVID-19 cases using a Luminex bead-based assay

**DOI:** 10.1101/2020.07.25.20161943

**Authors:** Joachim Mariën, Johan Michiels, Leo Heyndrickx, Karen Kerkhof, Nikki Foque, Marc-Alain Widdowson, Isabelle Desombere, Hilde Jansens, Marjan Van Esbroeck, Kevin K. Ariën

**Affiliations:** Outbreak Research Team, Institute of Tropical Medicine, Antwerp, Belgium; Virology Unit, Deparment of Biomedical Sciences, Institute of Tropical Medicine, Antwerp, Belgium; Department of Clinical Sciences Department, Institute of Tropical Medicine, Antwerp, Belgium; Immune response, Sciensano, Brussels, Belgium; University Hospital Antwerp, Antwerp, Belgium; University of Antwerp, Antwerp, Belgium

## Abstract

Large-scale serosurveillance of severe acute respiratory syndrome coronavirus type 2 (SARS-CoV-2) will only be possible if serological tests are sufficiently reliable, rapid and inexpensive. Current assays are either labour-intensive and require specialised facilities (e.g. virus neutralization assays), or expensive with suboptimal specificity (e.g. commercial ELISAs). Bead-based assays offer a cost-effective alternative and allow for multiplexing to test for antibodies of other pathogens. Here, we compare the performance of four antigens for the detection of SARS-CoV-2 specific IgG antibodies in a panel of sera that includes both severe (*n*=40) and mild (*n*=52) cases, using a neutralization and a Luminex bead-based assay. While we show that neutralising antibody levels are significantly lower in mild than in severe cases, we demonstrate that a combination of recombinant nucleocapsid protein (NP), receptor-binding domain (RBD) and the whole spike protein (S1S2) results in a highly sensitive (96%) and specific (99%) bead-based assay that can detect IgG antibodies in both groups. Although S1-specific IgG levels correlate most strongly with neutralizing antibody levels, they fall below the detection threshold in 10% of the cases in our Luminex assay. In conclusion, our data supports the use of RBD, NP and S1S2 for the development of SARS-CoV-2 serological bead-based assays. Finally, we argue that low antibody levels in mild/asymptomatic cases might complicate the epidemiological assessment of large-scale surveillance studies.

## Introduction

Since December 2019, Severe acute respiratory syndrome coronavirus type 2 (SARS-CoV-2) has spread at an unprecedented speed and scale, resulting in 500,000 deaths, 11,500,000 diagnosed cases (as of 10/7/2020)^1^ and likely many more undiagnosed cases with mild or no symptoms (10 fold)^2^. Nucleic acid (PCR) tests are widely used for clinical diagnosis and provide good estimates on the duration of viral shedding^3–5^. However, it remains uncertain how many people develop protective antibodies after recovery and for how long^6–8^. Serological data is typically used to determine the overall attack rate and level of herd immunity in a given population, and lays the foundation for control and prevention policies.

Serological tests exist in a variety of different formats but not all are appropriate for large-scale serosurveillance^6–9^. The gold standard for serological testing remains the virus neutralization test (VNT)^7^. This assay format is very specific and directly assesses the neutralizing capacity of antibodies in serum^10^. However, VNTs are also labour-intensive and generally require highly trained staff to work in BSL3 laboratory conditions. In contrast, enzyme-linked immunosorbent assays (ELISAs) require less trained operators and allow high-throughput screening, but are usually less specific due to cross-reactivity with other coronaviruses or other pathogens. Furthermore, ELISAs are relatively expensive since sufficient recombinant antigens needs to be produced^11^. Nevertheless, many commercial ELISAs became rapidly CE-labelled and are currently used for serosurveillance studies^7^. Since ELISAs as well as VNTs have important limitations for large-scale serosurveillance, microsphere bead-based assays using the Xmap Luminex technology have been increasingly developped^12–14^. This high-throughput platform allows the simultaneous detection of antibodies against different antigens from SARS-CoV-2, which can significantly increase the specificity of serological testing, in contrast to ELISA that usually includes only one antigen. Additional advantages of the bead-based assays are the need for lower serum amounts (<1µl) and the lower cost (as less recombinant antigen is required)^15^.

Evaluating a set of compatible immunogenic antigens is essential for the development of these multiplex bead-based assays. For many SARS-CoV-2 antibody tests, the main targets are the large spike glycoprotein (S) and the nucleocapsid protein (NP)^16^. The S protein is a trimeric class I fusion protein that consists of two subunits, namely S1 and S2^17,18^. The S1 protein mediates binding to host cells via interactions with the human receptor angiotensin converting enzyme 2 (ACE2) and is very immunogenic with its receptor-binding domain (RBD) as the main target for neutralizing antibodies^19^. The S2 subunit regulates fusion of the viral and host cellular membrane. The S protein, therefore, is an important target for the development of medical treatments and vaccines due to its role in cell binding and entry^20^. The NP plays a vital role in the transcription and replication of the virus, and is suggested to be more sensitive than the S protein for detecting early infections^16,21^. Here, we evaluate the performance of these four antigens (NP, RBD, S1, S1S2; and combinations between them) for the detection of SARS-CoV-2 IgG antibodies in a bead-based assay using sera from severe and mild cases in the early convalescent phase (<6weeks). We also examine the correlations between levels of SARS-CoV-2 neutralizing antibodies and those of antigen-binding antibodies measured by our Luminex assay and the commercial IgG/IgA Euroimmun ELISA.

## Methods

### Human serum samples

We used a panel of 286 serum samples: 40 samples from 22 hospitalized COVID-19 patients (severe cases) at the University Hospital of Antwerp (UZA) and confirmed by RT-qPCR, 52 samples from COVID-19 positive healthcare workers confirmed to have antibodies by VNT (mild/asymptomatic cases), and 191 leftover samples from the travel clinic of the Institute of Tropical Medicine (ITM) in Antwerp that we expect to be antibody negative because they were sampled in November 2019 (negative cases). Samples from severe cases were collected between March,3^th^ and April, 6^th^ from RT-qPCR confirmed COVID-19 cases (median days post symptom onset = 17, **Supplementary fig 1**). Of these, 17 severe cases were sampled two or three times over a period of one or two weeks. Samples from healthcare workers were collected between April,22^th^ and April,26^th^ in 17 different hospitals in Belgium. From this group, 45 had mild symptoms (median days post symptom onset = 25, **Supplementary fig 1)**, four were asymptomatic and six had an unknown symptom status. At the time of serum sampling, health care workers were also checked for viral RNA using a nasopharyngeal swap (two were RT-qPCR positive)^4^. All serum samples from severe and mild/asymptomatic cases underwent a viral inactivation protocol by heating at 56°C for 30 minutes.

### SARS-CoV-2 neutralization test

Serial dilutions of heat-inactivated serum (1/50-1/1600 dilutions in assay medium consisting of EMEM supplemented with 2 mM L-glutamine, 100 U/ml - 100 µg/ml of Penicillin-Streptomycin and 2% foetal bovine serum) were incubated with 3xTCID_100_ of a primary isolate of SARS-CoV-2 during 1h (37°C / 7% CO_2_). Sample-virus mixtures and virus/cell controls were added to Vero cells (18.000cells/well) in a 96well plate and incubated for 5 days (37°C / 7% CO_2_). The cytopathic effect caused by viral growth was scored microscopically. The Reed-Muench method was used to calculate the neutralising antibody titre that reduced the number of infected wells by 50% (NT50) or 90% (NT90)^22^, these values were used as proxy for the neutralizing antibody concentration in each sample.

### Luminex bead-based IgG immunoassay

We ordered commercially available recombinant NP, RBD, S1 and the complete Spike unit (S1S2) antigens derived from SARS-CoV-2 at Sino Biological (BIOCONNECT, Huissen, The Netherlands). The lyophilized proteins were resuspended in a buffer according to the manufacturer’s instructions and stored until use. Each antigens was coupled to a maximum of 10^6^ paramagnetic MAGPLEX COOH-microspheres beads from Luminex Corporation (Austin, TX), as described previously^15,23^. Each antigen was coupled at a concentration of 3µg per antigen to 1 × 10^6^ beads. Bovine Serum albumin (BSA, Sigma-Aldrich, St Louis, USA, 3µg) was coupled to an additional set of beads to serve as background control.

A microsphere working mixture was prepared in a phosphate buffered saline-BSA (PBS-BN) solution with a concentration of 2,000 beads/antigen/well. To choose an appropriate serum concentration, IgG titers were measured in serial dilutions (1/200-1/3,000) for five positive and three negative samples. We observed that a 1/1000 serum dilution gave the best signal to noise ratio (**Supplementary Fig 2**). Beads and diluted sera (1/1000) were added to each well in a final volume of 100µl. Plates were incubated at room temperature for two hours in the dark, and then washed three times with 200 µl/well of 0.05% PBS-Tween20. Subsequently, 100µl of a 1:400 secondary antibody (R-phycoerythrin-conjugated AffiniPure F(ab′)2 fragment of goat anti-human IgG, Jackson Immuno Research Laboratories) dilution in PBS-BN was added to each well. Plates were incubated for 45min in the dark at room temperature on a plate shaker (600rpm) and washed twice with PBS-BN-Tween. Beads were resuspended in 100µl PBS-BN and read using a Luminex^®^ 100/200 analyzer with 50 µl acquisition volume per well, DD gat 5000-25000 settings, and high PMT option. Results were expressed as crude median fluorescent intensities (MFI). The standard operating procedure for the bead-coupling and the assay is included in the supplementary material (SOP_Luminex).

### EUROIMMUN IgG/IgA ELISA

Samples from hospitalized cases were also analysed for level of antigen-binding antibodies against SARS-CoV-2 using the Anti-SARS-CoV-2 ELISA (IgA) and Anti-SARS-CoV-2 ELISA (IgG) (Euroimmun, Lübeck, Germany) following the manufacturer’s instructions. A ratio (optical density of the sample divided by optical density of the cut-off calibrator) <0.8 is considered negative, ≥0.8 and <1.1 borderline, and ≥1.1 positive^7^. The IgG and IgA Euroimmun ELISAs use the S1 spike protein, allowing comparison with S1-antibody levels of our Luminex assay.

### Statistical evaluation and diagnostic performance

To correct for background noise, we first subtracted the BSA-coupled bead signal (MFI_BSA_) from the median antigen-coupled bead signal (MFI_ag_), resulting in ΔMFI for each serum sample. We then investigated if mean ΔMFI levels (NP, RBD, S1, S1S2) differ significantly between serum of COVID-19 positive and negative cases using a student’s t-test. Subsequently, we tested if ΔMFI and neutralizing antibody levels differed significantly between serum of severe and mild cases using linear models. Because samples from the healthcare worker group were taken on average eight days later after symptom onset than samples from hospitalized patients (t=4.3781, df = 77.962, p = <0.0001, **Supplementary Fig 1**), we corrected for a potential sampling time effect by including days post symptom onset (dpo) in interaction with symptom status (severe vs mild). Neutralizing antibody levels were log-transformed to meet normality assumptions. When fitting the linear models, we started with the fully parameterised model (interaction between both independent fixed variables) and sequentially dropped variables that had the highest insignificant p-values. Significance was tested with the likelihood ratio test (assuming a *χ* ^2^ residual distribution). Furthermore, to test if antibody levels increase and stabilize for the severe group, we plotted individual ΔMFI values and neutralizing antibody levels for patients that were repeatedly sampled in function of dpo.

We assessed the sensitivity and specificity of the Luminex bead-based assay by looking at Receiver Operating Characteristics (ROC) curves. First, sensitivity and specificity were traded-off to calculate cut-off values for SARS-CoV2 antigens in singleplex. To test which combinations of antigens could improve the accuracy of the test in multiplex, we checked ROC curves calculated by supervised machine learning Random Forest (RF) algorithm models, as implemented in the R-package ‘randomForest’^24^. Variable (antigen) importance was assessed using the ‘varImplot’ function of the same package. Antigens with the lowest ‘mean decrease in accuracy’ and ‘mean decrease in Gini’ were sequentially removed from the bead set. Selection of optimal antigen combinations was performed based on the Area Under a Curve (AUC) values.

To assess the relationship between the different antigens and the neutralizing antibody response, we correlated the NT50 and NT90 measurements to ΔMFI and ELISA calibrator ratios using a Spearman’s rank (rs) test. To compare if beads coated with S1 antigen can be used as an alternative for the EUROIMMUN ELISA S1, we also correlated ELISA calibrator ratios to ΔMFI_S1_ values. All analyses were performed in the statistical software R.3.6.1.

## Results

### Differences in antibody levels between severe and mild cases

We first investigated whether ΔMFI levels for antigens (NP, RBD, S1, S1S2) differ between severe cases and mild cases, and the negative controls. The ΔMFI was significantly higher (p<0.0001) for positive than negative cases for all antigens (**Supplementary Fig 1**), indicating that antigen-coated beads can be used to discriminate positive from negative sera in singleplex. Neutralizing antibody (NT50 and NT90) and ΔMFI levels were also significantly higher for severe than for mild cases, although the significance level for ΔMFI_NP_ lay just below the 0.05 cut-off value (**Table 1; Fig 1**). A significant increase of antibody level in function of dpo was only observed for the NT50 and NT90 in the severe group (p-value=0.02), which were on average sampled eight days earlier than the mild ones (**Fig 2**). Consequently, neutralizing antibody and ΔMFI levels increased or remained constant for the majority of severe cases (*n*=16/18) that were repeatedly sampled 5-36dpo (**Supplementary Fig 3 and 4**).

**Table 1:** P-values expressing the effects of disease severity (severe vs mild) on antibody levels as measured by the Luminex or VNT corrected for days post symptom onset (DPO)

| Test | Antigen/NT | DPO | Severity | DPO x Severity |
| --- | --- | --- | --- | --- |
| Luminex | NP | 0.34 | 0.04 | 0.35 |
|  | RBD | 0.76 | 0.01 | 0.52 |
|  | S1 | 0.28 | <0.0001 | 0.71 |
|  | S1S2 | 0.83 | <0.0001 | 0.78 |
| Neutralization | Log(NT50) | Interaction | <0.0001 | 0.02 |
|  | Log(NT90) | Interaction | <0.0001 | 0.02 |

**Fig 1:**
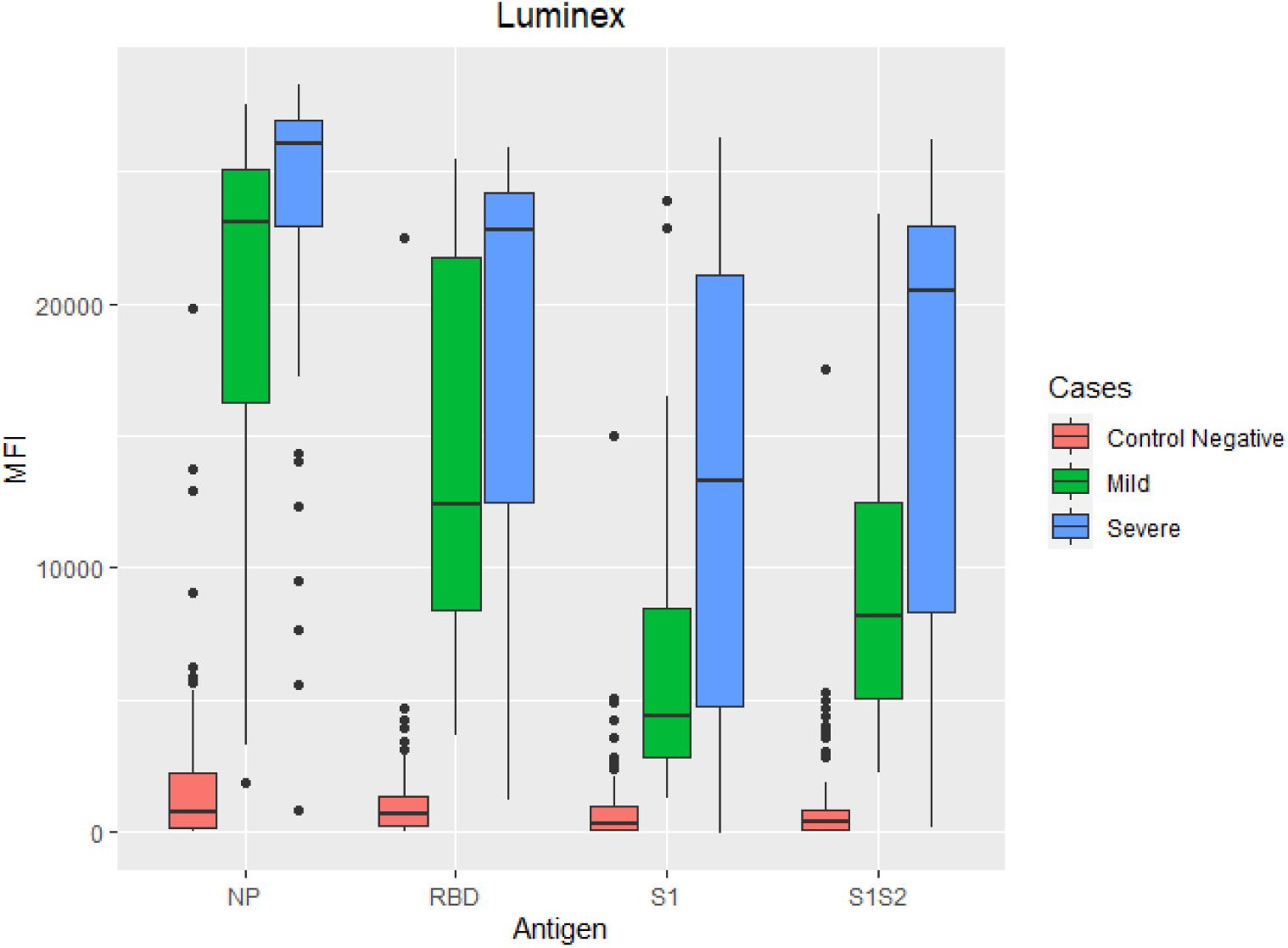
Boxplots representing differences in ΔMFI levels for NP, RBD, S1 and S1S2 between the severe (*n*=40, blue), mild cases (*n*=52, green) and the negative control (*n*=191, red) group.

**Fig2:**
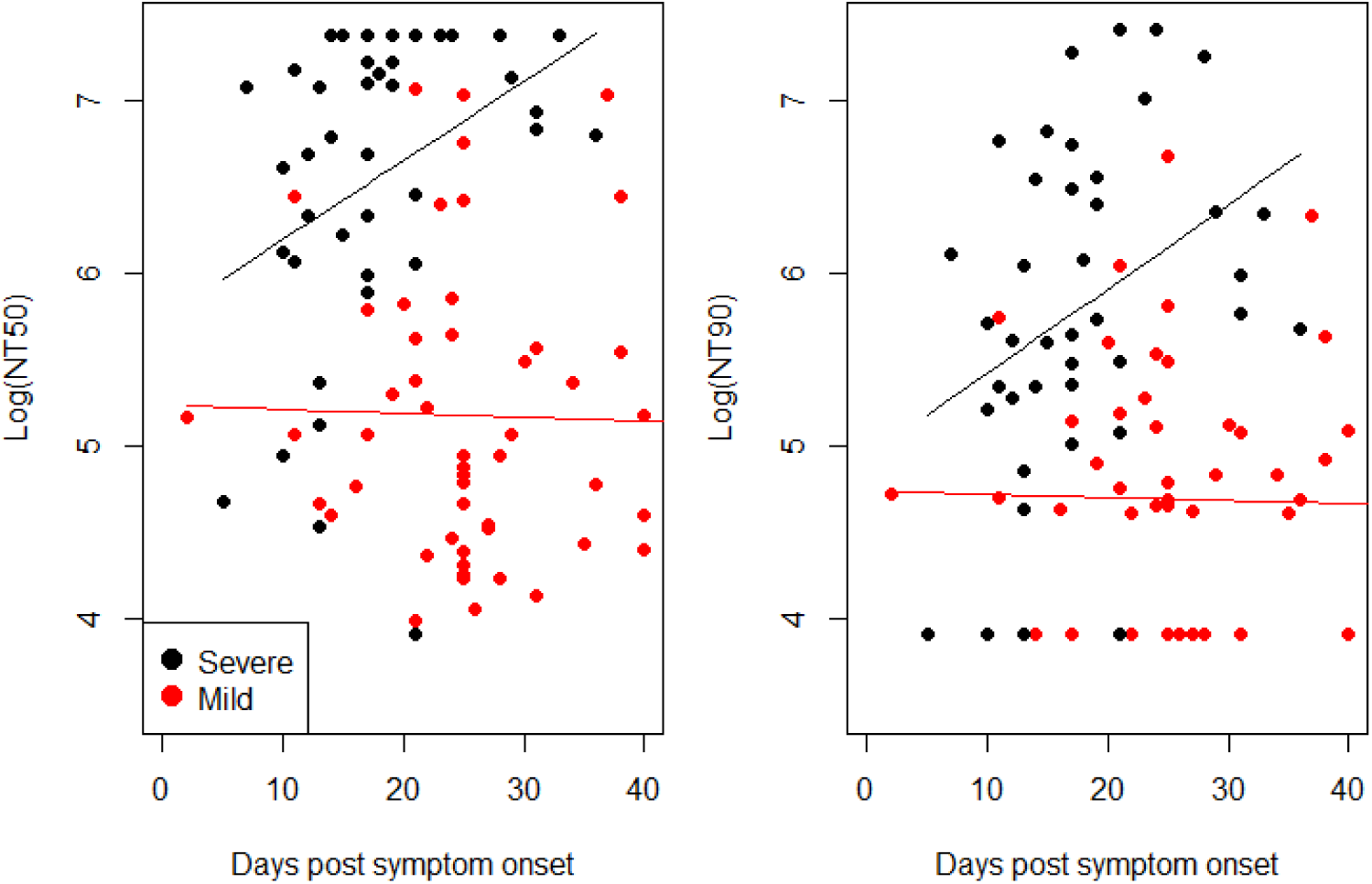
Comparison between log-transformed neutralizing antibody titers (NT50 and NT90) for severe (black) and mild cases (red) corrected for days post symptoms onset. Lines indicate the mean antibody level increase in function of dpo as predicted by the linear models.

### Performance assessment of the Luminex bead-based assay

Sensitivity, specificity and cut-off values were first calculated for individual antigens using a ROC analysis (**Fig 3** and **Table 2**). The large variation in AUC values suggests significant differences in classification performance for each antigen in singleplex. RBD had the highest classification performance (AUC=0.97) and S1 the lowest (AUC=0.83), while S1S2 (AUC=0.95) and NP (AUC=0.94) had almost the same classification performance. Subsequently, we calculated cut-off levels for each antigen by balancing sensitivity and specificity so that both were approximately equal. Similarly, the highest specificity and sensitivity were obtained for RBD and the lowest for S1. Final ΔMFI cut-off levels were: 3531 (RBD), 3621 (S1S2), 5507 (NP) and 1835 (S1) (**Fig 4**).

**Table 2:** Sensitivity and specificity of the Luminex antibody assay for antigens in single- and multiplex.

| Antigen(s) | AUC | sensitivity (%) | specificity (%) | cut-off (ΔMFI)/<br>random forest |
| --- | --- | --- | --- | --- |
| NP-RBD-S1-S1S2 | 0.993 | 96 | 99 | Random forest |
| NP-RBD-S1S2 | 0.993 | 96 | 99 | Random forest |
| NP-RBD | 0.992 | 96 | 97 | Random forest |
| RBD | 0.971 | 96 | 97 | 3531 |
| S1S2 | 0.949 | 94 | 97 | 3621 |
| NP | 0.940 | 96 | 96 | 5507 |
| S1 | 0.832 | 89 | 91 | 1835 |

**Fig 3:**
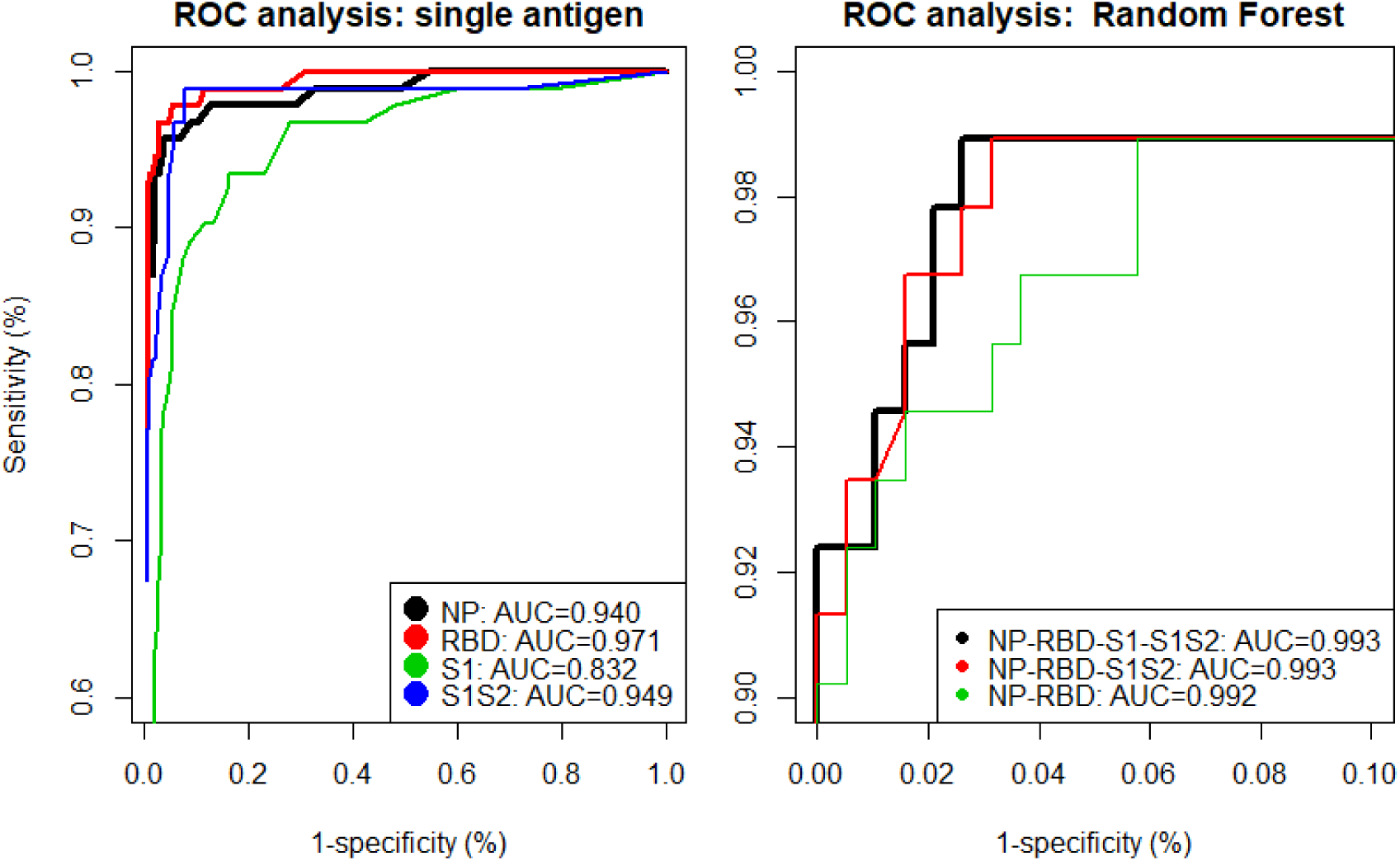
Receiver Operating Characteristic (ROC) curves obtained by varying the ΔMFI cut-off levels for antigens in singleplex (left figure); or combinations of antigens in multiplex as calculated by a random forest algorithm (right figure). The area under the curve (AUC) was used to evaluate the antigen’s classification performance.

**Fig 4:**
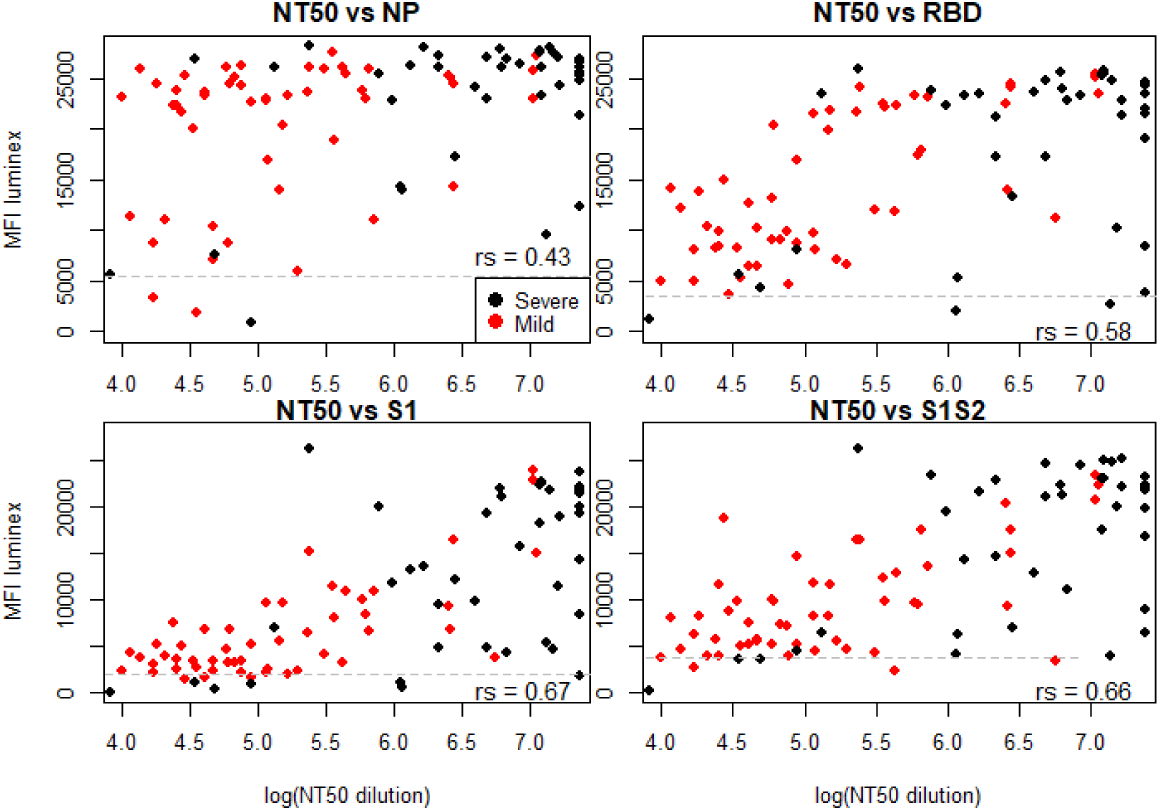
Correlations between NT50 and ΔMFI levels to NP, RBD, S1 and S1S2. Red dots represent serum from mild and black dots serum from severe COVID-19 cases, as calculated by the nonparametric Spearman correlation test (rs). Seropositivity cut-off levels are indicated by the dashed grey lines.

Subsequently, we used a random forest algorithm to test if combinations of different antigens could improve the classification performance of the assay. While multiplexing all antigens gave the best specificity (99%), the removal of S1 did not change the AUC (0.993) and sensitivity (96%) while the specificity decreased by 0.3% only. The variable importance scores (‘mean decrease in accuracy’ and ‘mean decrease in Gini’) also suggested that S1 can be removed from the multiplex without serious reduction in classification performance (**Supplementary Fig 5**). If we removed S1S2 (second lowest variable importance score), the prediction performance of the model decreased by ΔAUC=0.001.

### Correlations between neutralizing antibody, ΔMFI and ELISA levels

To assess if ΔMFI and ELISA levels can be used as proxy for the neutralizing capacity of serum, we correlated these levels to the NT50 (**Fig 4**) and NT90 (**Supplementary Fig 6**). We found overall moderate to strong correlations between ΔMFI values from antigens belonging to the S-protein and the NT50 (rs=0.58-0.67, p<0.0001) and NT90 (rs =0.59-0.68, p<0.0001), but a rather weak correlation between ΔMFI_NP_ and NT50 (rs=0.43, p=0.0001) and NT90 (rs=0.41, p=0.0001). This pattern can be explained by the observation that ΔMFI_NP_ already reached its maximum (ΔMFI_max_ = 28.000) at low neutralizing levels, which was not the case for ΔMFI_S1_ and ΔMFI_S1S2_. We also found that neutralizing antibody levels correlated stronger to ΔMFI levels of mild than severe cases (**Supplementary table 1**). Interestingly, NT50 (ρ=0.71, p<0.0001) and NT90 (rs =0.73, p<0.0001) correlated most strongly with IgA ELISA ratios (**Fig 5**). As expected, levels of ΔMFI_S1_ and IgG ELISA ratios also correlated strongly (rs =0.73, p<0.0001, **S Fig 7**).

**Fig 5:**
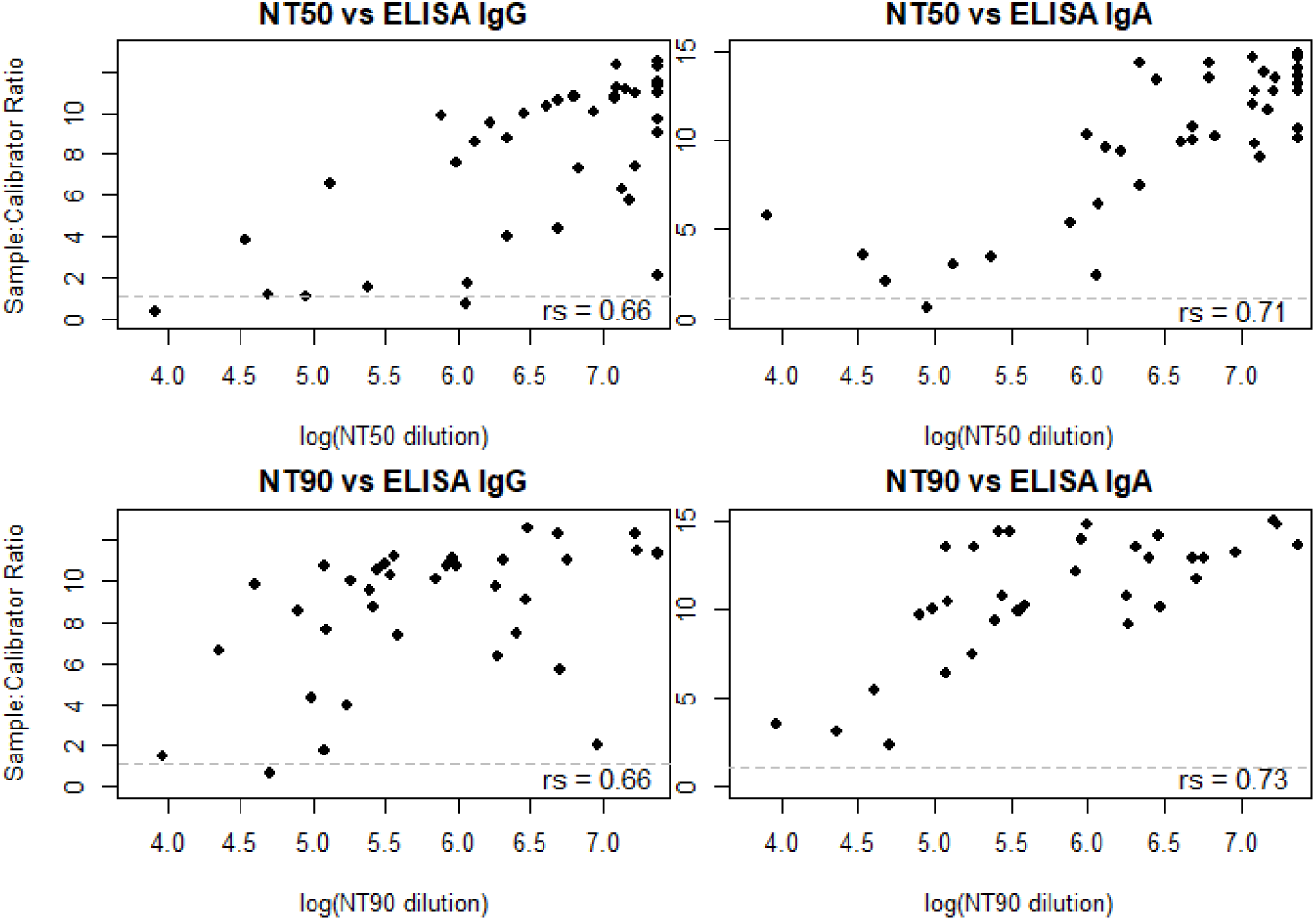
Correlations between the NT50/NT90 and Euroimmun IgG/ IgA ELISA ratios, as calculated by the nonparametric Spearman correlation test (rs). Seropositivity cut-off levels are indicated by the dashed grey lines.

## Discussion

While a plethora of commercial serological tests became available months after the discovery of SARS-CoV-2, researchers have already experienced problems in terms of sensitivity and specificity with many early market commercial tests^9^. Problems may arise because tests are mainly evaluated on sera from severe COVID-19 patients, who are suggested to develop a much higher immune response than mild or asymptomatic cases^25^. Here, we convincingly show that neutralizing and binding IgG antibody levels are indeed significantly higher for severe than mild/asymptomatic cases in the early convalescent phase (<6 weeks) (**Fig 1**). Furthermore, given that antibody levels directed to other human coronaviruses are known to decrease rapidly after infection (usually after 0.5-1 year)^26,27^ and seroreversions (antibody positive to negative) are already noted for asymptomatic SARS-CoV-2 infections^28^, we expect that antibody levels of the majority of mild/asymptomatic cases might drop below the cut-off value of many diagnostic tests within months to a year after infection. While we highlight that seroreversion does not necessarily mean that recovered people will become susceptible for COVID-19 months after the infection (immunity also depends on the T-cell mediated immune responses that might last much longer)^29^, it is clear that this issue will complicate the epidemiological assessment and should be taken into account by mathematical models that aim to understand/predict the transmission dynamics in the population^3,14^. This issue also raise questions related to vaccine development. Indeed, if humoral immunity would already decrease rapidly after natural infection, concerns exist that vaccines may not be able to trigger a sufficiently high and long-lasting protective antibody-mediated immune response ^30^.

The ROC analyses showed that NP, RBD and S1S2 antigens together can be used to develop a highly specific (99%) and sensitive (96%) Luminex SARS-CoV-2 antibody detection assay. Although these antigens performed well in singleplex (table 1), a combination of the three antigens clearly increased the assay’s prediction performance in comparison to the best performing singleplex (RBD, ΔAUC=0.022). The increase in specificity is explained by the multiplexing of three antigens in relation to the random forest algorithm. For example, a few aspecific bindings were noted in the negative control samples (4x NP and 1x RBD, S1 and S2) (**Fig 2**). While these samples would clearly be categorized as positives in a singleplex, they were actually categorized as negative by the random forest as the aspecific bindings did not happen in the same sample. This aspect makes a multiplex assay appealing for serosurveillance in sub-Saharan Africa, where malaria is notoriously known to cause false positive results in serological assays^31^. Interestingly, while RBD clearly outperformed the other antigens in a singleplex for classification (**Table 2**), antibody levels to NP were only slightly lower for severe than mild cases (**Fig 1**). This could be relevant in the light of a weaker immune response in mild/asymptomatic cases, suggesting that NP antibody levels might be easier to pick-up during serosurveillance studies (higher sensitivity)^21^, especially when overall antibody titers would decrease over time^27^. Consequently, a combination of NP with RBD/S1S2 is also put forward in two independent Luminex bead-based assays that were recently published ^12,13^.

Because VNTs are labour-intensive, require biosafety level-3 (BSL-3) conditions and specific training skills, simple serological tests that could predict individual levels of protecting- immunity are highly needed to assess the effect of vaccine campaigns^19^. For this reason, we correlated neutralizing titers (NT50 and NT90) to antibody levels obtained by our Luminex assay and a commercial ELISA. We found moderate-to-weak correlations to NP (rs_NT50_NP_=0.43; rs_NT90_NP_=0.41), probably because antibody levels already reached the maximum ΔMFI at low NT50 and NT90. The highest correlations were observed to the S1-antigen (rs_NT50_IgG:S1_=0.66; rs_NT90_IgG:S1_=0.68). Given that S1 is also the antigen target of the Euroimmun ELISA and our Luminex assay correlated strongly with the IgG ELISA in singleplex, it is no surprise that the IgG ELISA ratios correlate similarly with the neutralizing titers. While the inclusion of S1 to the Luminex bead set does not significantly improve the prediction performance of the test (**Table 2**), it can be included to better predict levels of neutralizing antibodies (this will only slightly increase the price per sample). Interestingly, the highest correlations were observed with the IgA ELISA (rs_NT50_ IgA:S1_=0.71; rs_NT90_ IgA:S1_=0.73), indicating that IgA might be a better predictor of neutralizing antibody levels than IgG during the early convalescent phase (<6 weeks).

In conclusion, we have developed a highly sensitive and specific serological assay for the detection of SARS-CoV- 2 IgG antibodies using a robust and high-throughput technology. Our assay can predict neutralizing antibody levels in the early convalescent phase relatively accurately. We also found that neutralizing titers and binding IgG antibodies differ significantly between severe and mild/asymptomatic COVID-19 cases, indicating that serological tests are best evaluated on serum panels that include mild and asymptomatic cases before use in large-scale serosurveillance settings.

## Data Availability

Data supporting the conclusions of this article are included in the article and its additional files. The datasets used and/or analysed during the current study are available from the corresponding author upon reasonable request.

## Ethics statement

Approval to sample from COVID-19 cases was obtained from the Ethical committee of the University of Ghent (BC-07587), local committees of each participating hospital and all participants provided consent to participate. The use of prepandemic leftover samples was approved by ITM’s internal review board. We declare that the planning conduct and reporting of the study was in line with the [Declaration of Helsinki](https://www.wma.net/policies-post/wma-declaration-of-helsinki-ethical-principles-for-medical-research-involving-human-subjects/), as revised in 2013.

## Acknowledgements

We thank Laure Mortgat, Elza Duysburgh, Natalie Fischer, Isabelle Thomas, Cyril Barbezange, Veronik Hutse for the organization of the Health Care Worker seroprevalence study (Sciensano/ITM, NCT04373889).

## Author contributions

Conceived the study: JMa, MAW and KA. Wrote the paper: JMa. Performed the lab experiments: JMa, JMi, LH, KK, and NF. Performed the statistical analyses: JMa. Supervised data collection and laboratory work: ID, HJ, MvE and KA. All authors read and approved the final manuscript.

## References

1. Johns Hopkins University. COVID-19 Dashboard by the Center for Systems Science and Engineering (CSSE). https://coronavirus.jhu.edu/map.html (2020). doi:10.1136/bmj.2.5361.864

2. Centers for disease control and prevention. Commercial Laboratory Seroprevalence Survey Data. https://www.cdc.gov/coronavirus/2019-ncov/cases-up https://www.cdc.gov/coronavirus/2019-ncov/cases-up (2020). Available at: https://www.cdc.gov/coronavirus/2019-ncov/cases-updates/commercial-lab-surveys.html.

3. Borremans, B. et al. Quantifying antibody kinetics and RNA shedding during early-phase SARS-CoV-2 infection. medRxiv 10.1101/2020.05.15.20103275 (2020). doi:10.1101/2020.05.15.20103275

4. Corman, V. M. et al. Detection of 2019 novel coronavirus (2019-nCoV) by real-time RT-PCR. Euro Surveill. 25, 1–8 (2020).

5. Hu, Z. et al. Clinical characteristics of 24 asymptomatic infections with COVID-19 screened among close contacts in Nanjing, China. Sci. China Life Sci. 63, 706–711 (2020).

6. Zhang, W. et al. Molecular and serological investigation of 2019-nCoV infected patients: implication of multiple shedding routes. Emerg. Microbes Infect. 9, 386–389 (2020).

7. Lassaunière, R. et al. Evaluation of nine commercial SARS-CoV-2 immunoassays. medRxiv 1–15 (2020).

8. Okba, N. M. A. et al. SARS-CoV-2 specific antibody responses in COVID-19 patients.medRxiv (2020). doi:10.1101/2020.03.18.20038059

9. Krammer, F. & Simon, V. Serology assays to manage COVID-19. Science 1227, 1–5 (2020).

10. Xun, J. et al. Neutralizing antibody responses to SARS-CoV-2 in a COVID-19 recovered 2 patient cohort and their implications. The lancet (preprint) (2020).

11. Amanat, F. et al. A serological assay to detect SARS-CoV-2 seroconversion in humans.Nat. Med. (2020). doi:10.1101/2020.03.17.20037713

12. Dobaño, C. et al. Highly sensitive and specific multiplex antibody assays to quantify immunoglobulins M, A and G against SARS-CoV-2 antigens. bioRxiv (2020). doi:10.1101/2020.06.11.147363

13. Ayouba, A. et al. Multiplex detection and dynamics of IgG antibodies to SARS-CoV2 and the highly pathogenic human Coronaviruses SARS-CoV and MERS-CoV. J. Clin. Virol. 104521 (2020). doi:10.1016/j.jcv.2020.104521

14. Rosado, J., Cockram, C., Merkling, S. H., Demeret, C. & Meola, A. Serological signatures of SARS-CoV-2 infection : Implications for antibody-based diagnostics. medRxiv 1–30 (2020).

15. Kerkhof, K. et al. Implementation and application of a multiplex assay to detect malaria-specific antibodies: a promising tool for assessing malaria transmission in Southeast Asian pre-elimination areas. Malar. J. 14, 1–14 (2015).

16. Taskin Tok, T., Tatar, G. & Tugba, T. T. Structures and Functions of Coronavirus Proteins: Molecular Modeling of Viral Nucleoprotein-International Journal of Virology & Infectious Diseases International Journal of Virology & Infectious Diseases. Int. J. Virol. Infect. Dis. 2, 1–7 (2017).

17. Wang, C. et al. A human monoclonal antibody blocking SARS-CoV-2 infection. Nat. Commun.11, 1–6 (2020).

18. Walls, A. C. et al. Structure, Function, and Antigenicity of the SARS-CoV-2 Spike Glycoprotein. Cell 181, 281–292.e6 (2020).

19. Premkumar, L. et al. The receptor binding domain of the viral spike protein is an immunodominant and highly specific target of antibodies in SARS-CoV-2 patients. Sci. Immunol.5, 1–9 (2020).

20. Padron-Regalado, E. Vaccines for SARS-CoV-2: Lessons from Other Coronavirus Strains. Infect.Dis. Ther. 9, 255–274 (2020).

21. Burbelo, P. D. et al. Detection of Nucleocapsid Antibody to SARS-CoV-2 is More Sensitive than Antibody to Spike Protein in COVID-19 Patients. arXiv 53, 1689–1699 (2015).

22. Reed, L. & Muench H., A. A simple method of estimating fifty per cent endpoints. Am J Hyg 27, 493–497 (1938).

23. Ambrosino, E. et al. A multiplex assay for the simultaneous detection of antibodies against 15 Plasmodium falciparum and Anopheles gambiae saliva antigens. Malar. J. 9, 1–12 (2010).

24. Breiman, L. & Cutler, A. Package ‘randomForest’. Cran Repos. (2018). doi:10.1023/A

25. Zhang, Z. et al. Early viral clearance and antibody kinetics of COVID-19 among asymptomatic carriers. medRxiv (2020).

26. Edridge, A. W. D. et al. Human coronavirus reinfection dynamics: lessons for SARS-CoV-2. Medrxiv 1–10 (2020). doi:10.1101/2020.05.11.20086439

27. Edridge, A. W. D. et al. Coronavirus protective immunity is short-lasting. medRxiv 2020.05.11.20086439 (2020). doi:10.1101/2020.05.11.20086439

28. Long, Q. X. et al. Clinical and immunological assessment of asymptomatic SARS-CoV-2 infections. Nat. Med. (2020). doi:10.1038/s41591-020-0965-6

29. Gutierrez, L., Beckford, J. & Alachkar, H. Deciphering the TCR Repertoire to Solve the COVID-19 Mystery. Trends Pharmacol. Sci. 1–13 (2020). doi:10.1016/j.tips.2020.06.001

30. Callaway, E. Coronavirus vaccines: five key questions as trials begin. Nature 579, 481 (2020).

31. Esbroeck, M., Meersman, K., Michiels, J., Ariën, K. & Bossche, D.Van den. Specificity of Zika virus ELISA: interference with malaria. Euro Surveill. 29, 252–262 (2016).

